# Accelerating Mental Health Precision Trial: An Effective Visualization-Driven Tool for Power and Sample Size Estimation in Biomarker-based Study Designs

**DOI:** 10.64898/2026.05.06.26352613

**Authors:** Desmond Zeya Chen, Aoqi Xie, Clement Ma

**Affiliations:** Centre for Addiction and Mental Health, Toronto, Ontario, Canada; Division of Biostatistics, Dalla Lana School of Public Health, University of Toronto, Toronto, Ontario, Canada

**Keywords:** Shiny App, Basket Trial, Interim Analysis, visualization, Biomarker-guided trial, Adaptive designs

## Abstract

Precision medicine has given rise to a spectrum of biomarker-guided trial designs, from simple enrichment and strategy designs to more complex adaptive frameworks. To address the need for user-friendly tools that span this spectrum, we developed a unified R Shiny platform that first implements three standard designs: the randomize-all design, the enrichment design, and the biomarker-strategy design, allowing researchers to perform power and sample size calculations under each framework with intuitive inputs and visual outputs. Building on this foundation, the platform further extends to support two-stage general randomized basket trial designs with interim analysis, which can be viewed as a generalization of the standard designs to multiple biomarker-defined subgroups. The tool was rigorously validated by comparison with established R pipelines and published formulas, and user testing confirmed its intuitive interface. By providing seamless integration from standard to advanced designs under a common input–output framework, our platform enables researchers to directly compare power and sample size requirements across different design choices using the same underlying assumptions. The result is a freely accessible tool offering effective visualizations for the full spectrum of biomarker-guided trial designs, available at https://ampt.obicloud.ca/. Future improvements may further expand the tool’s capabilities to accommodate the increasing complexity of trial designs needed by the research community.

## 1. Introduction

The era of precision medicine has brought biomarker-guided trial designs to the forefront of clinical research. At the foundational level, three standard designs are widely used: the randomize-all design (testing treatment efficacy in the overall population regardless of biomarker status), the enrichment design (restricting randomization to biomarker-positive individuals), and the biomarker-strategy design (comparing a biomarker-guided assignment against a biomarker-agnostic strategy)^1^. Building on these standard frameworks, the master protocol trial design has emerged as a natural generalization that innovatively aggregates the evaluation of multiple treatments or disease subtypes^2^. Its flexibility, adaptability, and effectiveness aided it gaining popularity since the onset of the millennium^3^. More specifically, the basket trial design is a commonly used form of a single master protocol that evaluates a single targeted therapy across multiple disease subtypes or biomarker-defined subgroups. After the first master protocol managed the Imatinib Target Exploration Consortium Study B2225 basket trial design^4,5^, there has been a growing number of such trials being implemented^6,7^. Basket trials with biomarker enrichment have pioneered new frontiers in personalized medicine and thoroughly revolutionized the randomized trial design^8^. Unlike existing approaches that assume equal basket sizes^9^, our tool relaxes this assumption^10^.

### 1.1 Study Objectives and Existing Work

We aim to provide trial researchers with support in proposal writing, trial monitoring, and trial execution. Majority of trial design software facilitate multiplicity through multi-arm multi-stage (MAMS) designs, while only a few explicitly address biomarker-based trial designs. Berry Consultants developed a commercial software called FACTS^11^ (the “Fixed and Adaptive Clinical Trial Simulator”), which offers simulation-based analysis with highly flexible options across different trial classes. FACTS can simulate basket trial designs with early futility and interim analysis. However, sample size estimation was not supported, as researchers must search through the range of sample sizes manually. Among open-source packages and pipelines, the Integrated Platform for Designing Clinical Trials (https://trialdesign.org) currently features two basket design trial R pipelines based on Bayesian framework. One of these proposes a calibration to the existing Bayesian hierarchical approach^12^, significantly reducing type I error rate inflation compared to the uncalibrated version. The other, a Bayesian latent subgroup design^13^, clusters cancer types in basket trials into responsive and non-responsive subgroups, resulting in higher power and controlled type I error rates compared to standard approaches. Additionally, a frequentist pipeline that analytically solves for early futility of baskets is available in the supplemental section of Chen et al^9^. Hence, to further enhance the software applicability and user experience, and to present a holistic elucidation of proposed trial design, we developed a web-based interactive R Shiny App^14,15^, to fill the methodological gap in general biomarker-based trial design^16^. Although this overview highlights several essential tools and pipelines, it is important to recognize that to our knowledge, AMPT (Accelerating Mental Health Precision Trials) is the first open-source platform that unifies standard and basket designs under a single interface.

### 1.2 Target Audiences and Application Scope

Our tool is designed to support randomize-all, enrichment, biomarker-strategy designs, and two-stage general randomized basket trials. These four designs cover the full spectrum of biomarker-guided trial planning, from simple efficacy testing to adaptive multi-subgroup evaluation. Unlike standard trials that focus on a single disease at a time, basket trials assess the efficacy of a single treatment across multiple homogenous diseases, significantly reducing the required sample size when comparing to operating individual trials. The two-stage setup adds flexibility by allowing unresponsive disease baskets to be pruned at the interim stage, reallocating the remaining recruitment quota to responsive baskets. This early futility check ensures that resources are not wasted, improving the overall efficiency of the trial.

The tool is particularly valuable for clinical trial researchers and biostatistical support teams due to its accessibility, intuitive usage, and convenience. Its user-friendly interface simplifies the complex process of designing and analyzing biomarker-guided trials, making it easier for researchers to implement these designs without needing extensive statistical training. For users who primarily work with standard biomarker-guided designs, the platform provides quick, dedicated calculators that avoid the complexity of adaptive features. For those requiring more flexible frameworks, the platform also supports basket trial designs that accommodate heterogeneous biomarker-defined subgroups within a single trial. The development of our platform, AMPT, was motivated by challenges in mental health research, however, the methods and tools presented here are general and applicable to any disease area requiring biomarker-guided trial designs. While basket designs have been particularly effective in oncology drug development, the AMPT platform is equally applicable to any setting where biomarker prevalence varies across subgroups and sample size planning must account for such heterogeneity, including rare diseases and precision medicine more broadly.

## 2. Implementation

### 2.1 Design Details for AMPT

#### Standard Biomarker-Guided Designs

AMPT supports three standard designs for biomarker-guided trials (Figure 1). Each design is implemented as a dedicated calculator module, providing immediate power and sample size estimates without requiring adaptive inputs.

1. **Randomize-All Design.** All eligible participants are randomized to treatment versus control regardless of their biomarker status. The trial tests whether the treatment is efficacious across all participants.
2. **Enrichment Design.** All eligible participants are first screened for the selected biomarker. Only participants who screen positive for the biomarker (M⁺) are randomized to treatment versus control. The trial tests whether the treatment is efficacious in M⁺ participants only.

**Figure 1:**
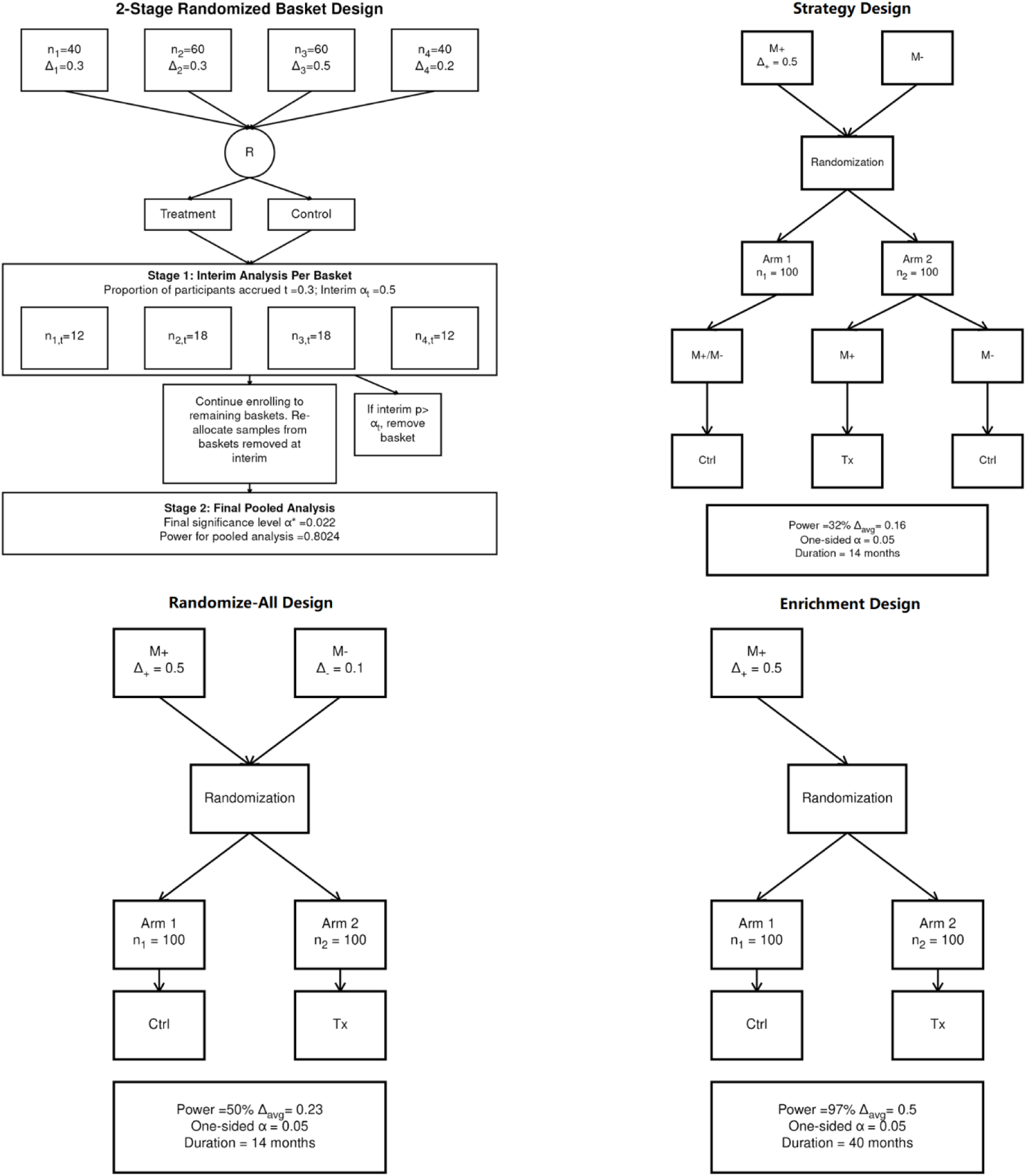
Study Schema of Each Biomarker-based Design Produced by AMPT

1. **Strategy Design.** All eligible participants are first screened for the selected biomarker. Participants are randomized to either a “biomarker-based strategy” (where M⁺ and M⁻ participants receive the experimental or control treatments, respectively) or to a “biomarker-agnostic strategy” (where all participants receive the control treatment). The trial tests whether using the biomarker-based strategy to guide treatment improves outcomes.

For each of these designs, power and sample size calculations follow standard formulas, with user inputs including expected effect sizes in M⁺ and M⁻ subgroups, biomarker prevalence, and type I error rate.

#### Two-Stage General Randomized Basket Trial Design

The basket trial design in AMPT follows the GRaBIt (Two Stage General Randomized Basket Trial Design) framework^10,17^ (Figure 1). Randomized basket trials test a common treatment versus control across several related disease subgroups (baskets). The GRaBIt design improves on prior basket methods by allowing each basket to have different sample sizes and treatment effects.

The trial proceeds in two stages:

- **Stage 1 (Interim).** A fraction of participants is enrolled and randomized in each basket. An interim analysis is performed to identify and remove baskets in which

the experimental treatment is not efficacious (based on a prespecified interim significance level).

- **Stage 2 (Final).** Participants in the remaining baskets are enrolled. A final pooled analysis is performed using Stage 1 and Stage 2 data from the remaining baskets only.

The null hypothesis is that the experimental treatment is not superior to control in any of the prespecified baskets; the alternative hypothesis assumes superiority in at least one basket. Following the statistical framework of Chen et al^9^ and Patel et al^10^, the overall Type I error rate is computed as the sum, under the null, of rejection probabilities for baskets that pass both the interim and final stages. Power is estimated similarly under the alternative, with the rejection region adjusted by a noncentrality parameter for active baskets.

To avoid algebraically solving the sample size formula directly, we use a root-finding approach based on the power function. Only the overall trial sample size needs to be determined; basket-specific sample sizes are derived from disease prevalence ratios. Because power increases monotonically with sample size, we implement a recursive binary search^18^. This algorithm is substantially faster than exhaustive iterative loops, directly improving the responsiveness of the Shiny application.

### 2.2 Variable Definitions and Input Parameters

Both calculations require the user to provide crucial trial characteristics (Figure 2).

**Figure 2:**
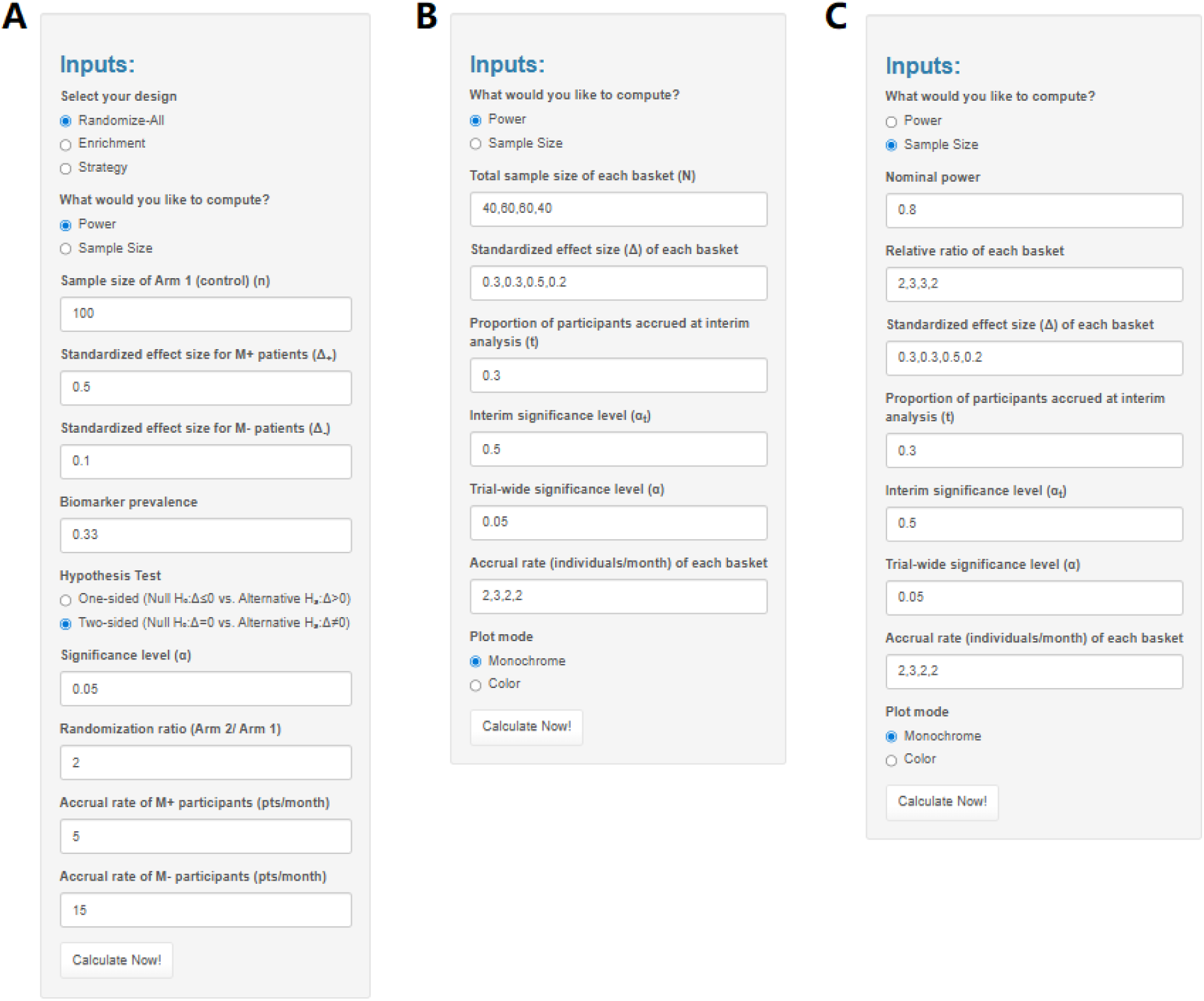
Input Parameters Required for Power or Sample Size Calculation of GRaBIt

For the standard designs (randomize-all, enrichment, and strategy), the standardized effect sizes for biomarker-positive (M⁺) and biomarker-negative (M⁻) patients, denoted Δ⁺ and Δ⁻, must be specified; investigators can estimate these values from published literature or pilot studies. The prevalence of the biomarker in the general population is also required. For hypothesis testing, the user selects either two-sided (null of equality) or one-sided (null of less-than). The significance level (α) must be provided, representing the overall false positive rate. The randomization ratio (allocation of experimental/strategy arm to control arm) and the accrual rates of M⁺ and M⁻ participants (in pts/month) are also required; the accrual rates primarily affect the trial duration estimation rather than the power or sample size calculations. For power calculation (Figure 2A), the user provides the sample size of arm 1; conversely, for sample size calculation, the user provides the targeted power (1−β), and the tool returns the required sample size of both arms.

For the GRaBIt basket trial design, the user provides similar but basket-specific inputs (Figure 2B). The standardized effect size (Δ) for each basket must be provided; investigators can estimate these values based on subtype prevalence from published results. The proportion of participants accrued at interim analysis (t) is also required. This is the cutoff point for interim analysis, expressed as a percentage of the total participants recruited, and should be consistent across all baskets. The interim significance level (αₜ) is required, which is the designated α level for the interim analysis at proportion t. Any basket with a P-value exceeding this threshold will be dropped. The trial-wide significance level (α) must also be provided.

For sample size calculation in the basket design, the sample size per basket is required (Figure 2C). Conversely, when calculating the sample size per basket, the nominal power (designated 1−β level for the entire trial, or one minus the expected type II error rate) and the relative ratio of sample size in each basket must be provided. These are the primary parameters that directly influence the calculation, while the number of individuals recruited per month in each basket only affects the trial duration estimation.

### 2.3 System Architecture and User Interface Design

Having cloud computing power freely provided by the R Shiny^15^ server, the local machine is only responsible for capturing inputs and rendering outputs on the user interface, requiring minimal hardware resources. The remote server handles heavy-duty statistical computing tasks. The main tab of the tool consists of two sections: the input panel and the output panel. Power and sample size calculations are unified through an initial question that asks users which type of calculation they intend the tool to perform. The corresponding trial characteristics are then required for the calculation of their choice. On the output panel, an interactive trial design plot is displayed above the text summary. If incorrect inputs are provided, error messages will appear, masking all outputs. Additionally, a’Readme’ tab is available, offering an example, detailed explanations of the interface, and the statistical methods used, helping users gain hands-on experience.

### 2.4 Operational Workflow and Error Detection

Users can input trial characteristics with just a few clicks and text entries. The app is initiated by clicking the’Calculate Now!’ button. All input parameters are sent to the backend, where they are checked for missing data or syntax errors before being passed into the function. The function then verifies that all input sample sizes are valid whole numbers. If any errors are found, they are promptly flagged for correction. Once all error checks are passed, the power or sample size is computed. Both summary statistics and trial characteristics inputs are compiled into a plot to visually represent the proposed trial.

A text summary of the plot is displayed below for additional clarity. Users could then download the plot and text summary separately for their intended use. Alternatively, the single-click export module compiles graphical outputs, summary tables, and parameter summaries into a pre-formatted Word template without requiring external software.

### 2.5 Validation, Reproducibility, and User Testing

To enhance reproducibility, we conducted extensive validation analysis and user testing. App’s outputs were systematically compared with results generated by the original R scripts used in the underlying statistical calculations. This cross-verification ensured that the app replicates the established methods accurately. A variety of edge cases were tested, including extreme values of parameters and atypical trial configurations, to confirm the app handles all inputs robustly and responsively. This is crucial for identifying any scenarios where the app will fail and imposing corresponding error checkers as countermeasures. Additionally, we carefully examined the app’s error reporting mechanisms to ensure they provide clear and actionable feedback to users. This included verifying that the app correctly identifies and flags incorrect or missing inputs before proceeding with calculations. Finally, we conducted user acceptance testing with potential end-users like clinicians and biostatisticians to validate the app’s functionality in real-world scenarios. Participants were asked to replicate an example provided in the questionnaire, specifically for testing reproducibility. Feedback from these tests was incorporated to refine the app’s interface and usability.

## 3. Results

Our web application for two-stage general basket trial design and standard biomarker-guided designs is hosted at https://ampt.obicloud.ca/. To ensure broad accessibility, the app is fully responsive, allowing seamless operation on desktops, tablets, or even smartphones. The intuitive user interface (Figure 3) is designed to cater to users with varying levels of statistical knowledge, ensuring that even those unfamiliar with advanced methods can easily navigate the tool. The interactive trial design figure can be downloaded as a JPEG file, and the design summary provides a clear, technical explanation of each trial characteristic along with summary statistics of the proposed design. A Word export button was provided, of which can automatically knit both figure and design summaries into a unified word document. Both GRaBIt and standard design module incorporated the export button functionality.

**Figure 3:**
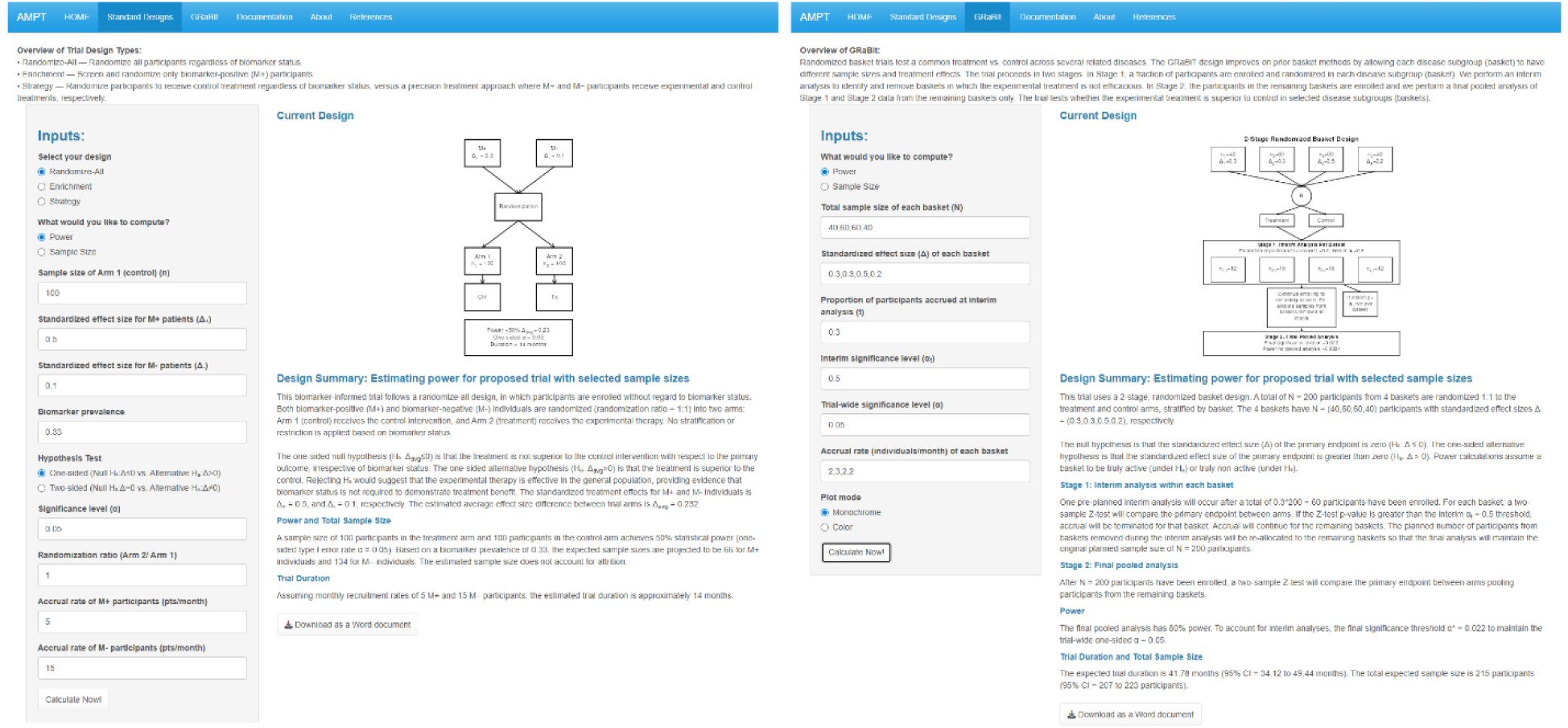
App Interface

### 3.1 User Interaction Feedback and Usability

Three users participated in the testing of the Shiny app, their feedback focused on ease of use, functionality, layout, and performance. All users found the app easy to navigate, especially with the help of default examples and detailed guidance provided in the Readme tab. Our original goal was to make the tool accessible to users with limited statistical training, aiming to simplify clinical trial design. However, one user noted that the use of highly specialized statistical language in the input variable names and explanations could create barriers for our target audience, potentially undermining our objective. We addressed this issue by incorporating additional explanations for each input from a trial design perspective, clarifying’What does this mean in clinical trial terminology?’ and’How could you decide on a value for it?’

The app was also presented at an internal meeting, where feedback related to sample size estimation functionality was raised. In response, we enhanced the app by adding features that support sample size estimation directly within the tool, further improving its utility for users designing clinical trials.

The app’s layout and functionality were deemed logical and well-organized, with users appreciating the clear instructions and intuitive design. Suggestions for enhanced result explanations and improved result visibility were carefully incorporated. In terms of performance, while the app generally worked well, users did experience occasional freezing or slower performance when working with larger sample sizes on the prototype version. These issues are common with the free version of the Shiny App server, and we have now migrated the App to an internal server hosted by the Ontario Brain Institute. All users expressed their willingness to use and recommend the app, citing its user-friendly interface and practical features. The user testing process, along with the feedback from the internal meeting, significantly improved the app’s usability and provided valuable, unbiased feedback from potential users.

### 3.2 Comparative Analysis for GRaBIt

Primary trial characteristics like the total sample size, interim sample size, and number of baskets are influenced by the interrelated inputs: the number of baskets, sample size per basket, and interim proportions. Selecting an interim alpha of 0.3 allows for effective interim analysis without being overly conservative during partial recruitment. The trial-wide alpha is set at the conventional two-sided significance level of 0.025. We validate the Shiny App by comparing it against an R pipeline, using selected combinations of these inputs while controlling for effect sizes and alphas (Table 1).

**Table 1:**
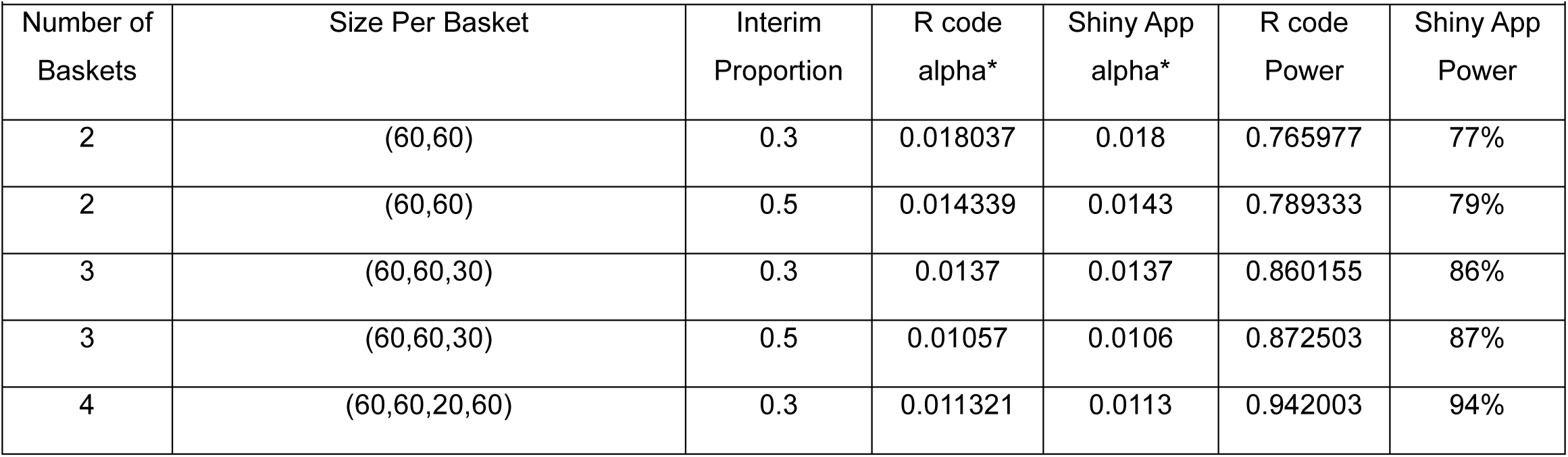
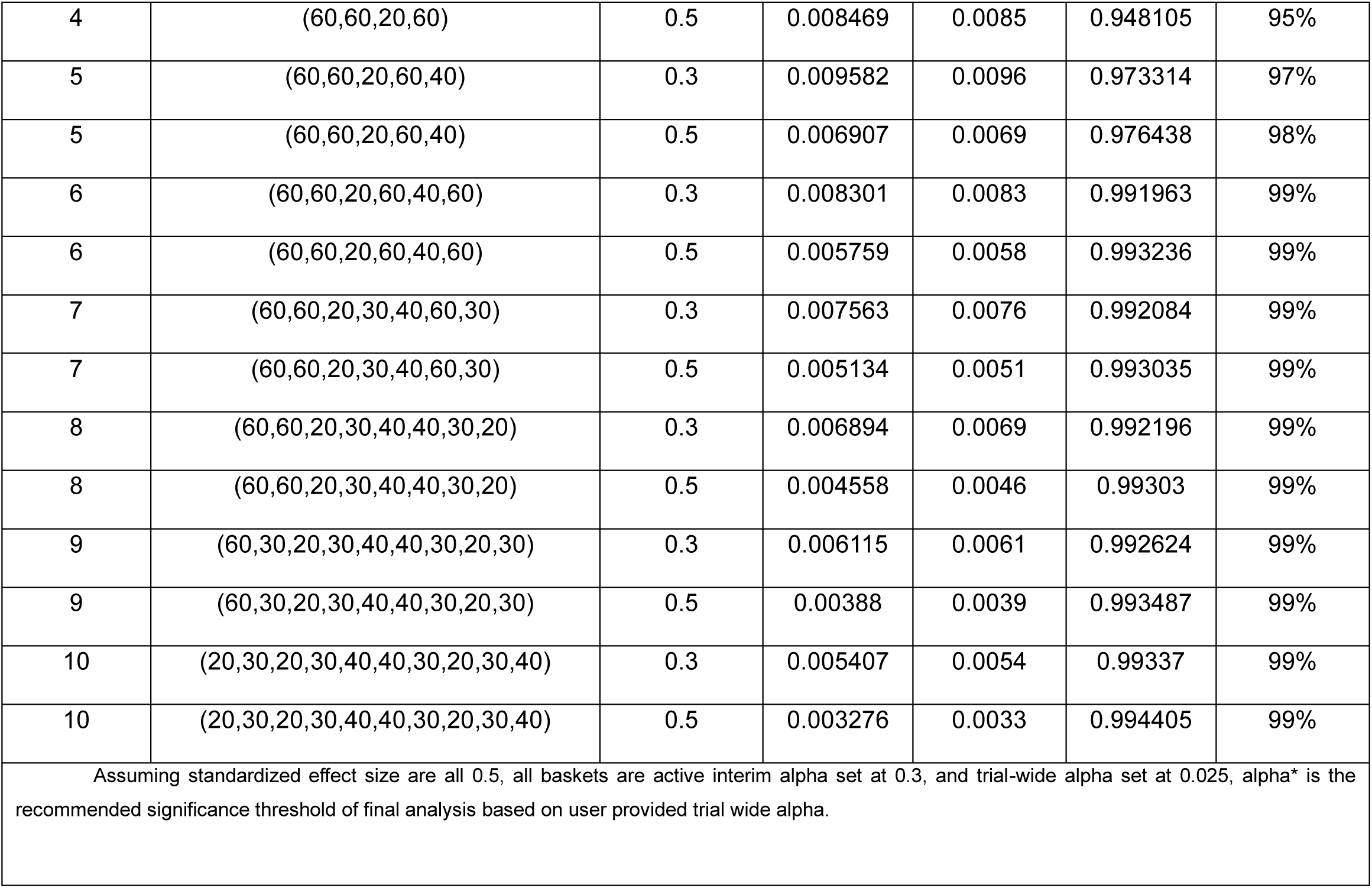
Comparative Analysis of the Tool vs R pipeline varying number of baskets and interim proportion.

The alternative hypothesis is defined by user-specified effect sizes, with a value of 0 indicating that a basket will be pruned during interim analysis. To validate the tool under different alternative hypothesis scenarios, we varied the effect sizes per basket while keeping the sample size, number of baskets, interim proportion, and alphas constant (Table 2).

**Table 2:** Comparative Analysis of the Tool vs R pipeline varying effect size per basket.

|  |  |  |  |  |  |
| --- | --- | --- | --- | --- | --- |
| Effect Size Per Basket | Baskets Remained at Interim | R code alpha* | Shiny App alpha* | R code Power | Shiny App Power |
| (0.2,0.2,0) | (1,1,0) | 0.01370037 | 0.0137 | 0.19221983 | 19% |
| (0.2,0.2,0.2) | (1,1,1) | 0.01369794 | 0.0137 | 0.23586268 | 24% |
| (0.5,0.2,0) | (1,1,0) | 0.01370189 | 0.0137 | 0.55850379 | 56% |
| (0.5,0.2,0.2) | (1,1,1) | 0.01369941 | 0.0137 | 0.58211621 | 58% |
| (0.5,0.5,0) | (1,1,0) | 0.01370145 | 0.0137 | 0.78413515 | 78% |
| (0.5,0.5,0.2) | (1,1,1) | 0.01370213 | 0.0137 | 0.79281031 | 79% |
| (0.5,0.5,0.5) | (1,1,1) | 0.0137017 | 0.0137 | 0.8601982 | 86% |
| Table 2: Varying effect size & whether the basket is truly active of (60,60,30). Interim proportion set at 0.3, interim alpha set at 0.3, and trial-wide alpha set at 0.025. Truly active vector, the corresponding baskets that pass interim analysis, deduced via effect size per basket that user provided. |  |  |  |  |  |

The results from the tool matched those generated by the R code after rounding, confirming that the tool correctly interprets the inputs and performs equivalently to the R code. This validation demonstrates the tool’s robustness and reliability in replicating established statistical processes. The consistency between the tool and the R code suggests that users can confidently employ the tool for interim analysis and hypothesis testing in clinical trials.

### 3.3 Comparative Analysis for Standard Biomarker-based Designs

Across all three standard biomarker-guided designs: enrichment, randomize-all, and strategy, the power values reported by the Shiny App matched those generated by the independent R pipeline after rounding (Table 3). This agreement held consistently across the full range of input configurations examined: two biomarker prevalence values (0.1 and 0.3), two randomization ratios (1:1 and 2:1), two Arm 1 sample sizes (100 and 200), and two standardized effect size pairs. For enrichment designs, power ranged from 56% to 100% in the Shiny App, corresponding to 0.5641 to 0.9999 in the R pipeline. For randomize-all designs, power ranged from 13% to 72% (0.1332–0.7192 in the R pipeline), and for strategy designs, from 4% to 41% (0.0402–0.4099 in the R pipeline). No discrepancies beyond rounding error were observed in any cell of the design space, confirming the tool’s correct implementation of the underlying computations and its reliability for designing biomarker-informed trials.

**Table 3:** Statistical Power of Biomarker-Guided Standard Designs: Shiny App vs R Code Validation.

|  |  |  |  |  |  |
| --- | --- | --- | --- | --- | --- |
| Biomarker Prevalence = 0.1 |  |  |  |  |  |
| Design | Source | 1:1 Randomization |  | 2:1 Randomization |  |
| | | $n_1=100$ | $n_1=200$ | $n_1=100$ | $n_1=200$ |
| Panel A: Effect size (0.3, 0.1) |  |  |  |  |  |
| Enrichment | Shiny | 56% | 85% | 69% | 93% |
|  | R code | 0.5641 | 0.8508 | 0.6878 | 0.9337 |
| Randomize-all | Shiny | 13% | 22% | 16% | 28% |
|  | R code | 0.1332 | 0.2236 | 0.1635 | 0.2829 |
| Strategy | Shiny | 4% | 5% | 4% | 5% |
|  | R code | 0.0402 | 0.0485 | 0.0432 | 0.0533 |
| Panel B: Effect size (0.5, 0.1) |  |  |  |  |  |
| Enrichment | Shiny | 94% | 100% | 98% | 100% |
|  | R code | 0.9424 | 0.9988 | 0.9831 | 0.9999 |
| Randomize-all | Shiny | 16% | 29% | 21% | 37% |
|  | R code | 0.1660 | 0.2878 | 0.2070 | 0.3657 |
| Strategy | Shiny | 5% | 7% | 6% | 8% |
|  | R code | 0.0541 | 0.0721 | 0.0604 | 0.0834 |
| Biomarker Prevalence = 0.3 |  |  |  |  |  |
| Design | Source | 1:1 Randomization |  | 2:1 Randomization |  |
| | | $n_1=100$ | $n_1=200$ | $n_1=100$ | $n_1=200$ |
| Panel C: Effect size (0.3, 0.1) |  |  |  |  |  |
| Enrichment | Shiny | 56% | 85% | 69% | 93% |
|  | R code | 0.5641 | 0.8508 | 0.6878 | 0.9337 |
| Randomize-all | Shiny | 20% | 36% | 26% | 46% |
|  | R code | 0.2037 | 0.3594 | 0.2567 | 0.4552 |
| Strategy | Shiny | 9% | 14% | 11% | 18% |
|  | R code | 0.0928 | 0.1446 | 0.1103 | 0.1786 |

| Panel D: Effect size (0.5, 0.1) |  |  |  |  |  |
| --- | --- | --- | --- | --- | --- |
| Enrichment | Shiny | 94% | 100% | 98% | 100% |
|  | R code | 0.9424 | 0.9988 | 0.9831 | 0.9999 |
| Randomize-all | Shiny | 34% | 59% | 43% | 72% |
|  | R code | 0.3430 | 0.5948 | 0.4350 | 0.7192 |
| Strategy | Shiny | 18% | 32% | 23% | 41% |
|  | R code | 0.1842 | 0.3228 | 0.2311 | 0.4099 |
| Table 3: Column $n_1$ refers to sample size in Arm 1 under a given randomization ratio. Standardized effect sizes ($\Delta_{m+}$, $\Delta_{m-}$) represent biomarker-positive and biomarker-negative subgroups, respectively. | | | | | |

### 3.4 Worked Example

Recent findings have established a link between biological sex differences in the FGF13 gene on the X chromosome and the development of Autism^19,20^. We hypothesize that biological sex differences in the FGF13 gene on the X chromosome may influence treatment response in Autism. To mimic this scenario, we simulate a study cohort divided into four groups: FGF13+ Males, FGF13- Males, FGF13+ Females, and FGF13- Females. Reflecting the potential for varying prevalence of FGF13+ mutations across sexes, we assign the following hypothetical group proportions: 30%, 30%, 10%, and 30%, respectively. Assuming a total sample size of 200, a 30% accrual rate, and an interim alpha of 0.3 for an interim analysis, this simulated trial would have a power of 94%. To maintain an overall Type I error rate of 0.025, the final significance threshold would be recommended to be 0.011 (Table 1).

## 4. Discussion

Our open-source calculator is compiled to run on browser while computing were done remotely at RShiny^15^ server. Both power and sample size calculation functions were integrated into the same interface. Instead of manually constructing the pipeline, calibrating inputs, rephrasing outputs so its human readable, and plotting the design, arranging plot and textual output into a word document; our visualization tool automates all above tasks with just few simple steps. It provides users with an integrated, statistics enriched trial design figure and detailed textual exposition of the trial design alongside statistical results. These outputs can be conveniently assimilated as presented.

This user-friendly approach not only saves time but also reduces the potential for human error in trial design, ensuring a higher degree of accuracy and consistency. The tool’s ability to produce ready-to-assimilate outputs enhances its practical value, allowing for immediate integration into trial planning and reporting.

As clinical research moves towards more complex trial designs, basket trials have become increasingly popular due to their efficiency in exploring the effects of interventions across multiple disease types. These designs allow clinicians to investigate multiple patient populations simultaneously, improving resource allocation and potentially accelerating the discovery of effective treatments. Our tool fits seamlessly into this evolving landscape, providing an accessible means for researchers to engage with these complex designs without the need for extensive statistical training. By supporting the development and execution of biomarker-based designs, the tool also holds potential for educational purposes, enabling clinicians and researchers to independently explore and visualize these innovative trial structures. This flexibility, combined with the potential to expand into multi-stage and multi-arm trial designs, pinpoints the importance of developing tools that can keep pace with modern research needs.

As biomarker-based designs gain popularity for biomarker-informed treatment outcome analysis, the sex variable, could either interact with, or directly affecting biomarkers intensity to influence efficacy^21,22^. Despite the prominence of biological sex-differentiated disease etiology in medical research^23^, limited attention has been given to incorporating the sex variable as the primary basis for basket grouping or post-trial outcome analysis^24^. We presented an emulating example of how basket trial could be utilized to disentangle the sex varied therapeutic effect.

Additionally, the Shiny application is natively compatible with multimodal AI agents capable of visual understanding and browser automation. Because the tool exposes a standard HTML interface, large language models can interact with it directly. A clinical investigator could describe a trial design in natural language to an AI assistant, and the assistant can takeover all remaining execution level tasks and return the investigator with a study summary. The trial design document is generated from stable, pre-authored templates reviewed by senior biostatisticians with trial design experience. The application performs deterministic parameter substitution into these templates to ensure its consistency and reproducibility; it does not rely on a language model to generate or interpret statistical content. The AI agent in this case serves solely as an intelligent input layer, translating natural language into precise UI operations.

In a nutshell, as the use of biomarker-based trials continues to expand, our tool is positioned to evolve alongside these developments, with potential future enhancements to support multi-stage designs, multi-arm trials, and other innovative trial methodologies. Gathering user feedback will be critical in guiding these developments to ensure the tool continues to meet the evolving needs of the research community.

## Author Contributions

D.Z.C. and C.M. conceptualized the study. C.M. supervised the study. D.Z.C. and C.M. drafted the manuscript. D.Z.C. performed the analyses, developed the visualization software, and summarized the results. A.X. contributed to the investigation by conducting user testing of the application.

## Declaration of interests

The author(s) declare no conflicts of interest.

## Data Availability

Not Applicable

## Acknowledgements

We would like to thank Marcos Sanches and the rest of the biostatistics core at CAMH for their insightful questions during the internal meeting, which played a key role in shaping the development of this application. We would like to acknowledge the support of the University of Toronto Fellowship, provided by the University of Toronto for funding this research, and generous funding by the Ontario Brain Institute Centre for Analytics and the CAMH Discovery Fund.

## Software and Web Resources

The visualization tool was developed using R^14^ version 4.3.2. The implementation involved several R packages, including shiny^15^ (version 1.8.0), shinythemes^25^ (version 1.2.0), ggplot2^26^ (version 3.4.4), ggtext^27^ (version 0.1.2), and ggforce^28^ (version 0.4.1).

## Notes

### Competing Interest Statement

The authors have declared no competing interest.

